# Elevated levels of IL-6 in IgA nephropathy patients are induced by an epigenetically driven mechanism modulated by viral and bacterial RNA

**DOI:** 10.1101/2022.03.05.22271944

**Authors:** Fabio Sallustio, Angela Picerno, Maria Teresa Cimmarusti, Francesca Montenegro, Claudia Curci, Giuseppe De Palma, Carmen Sivo, Francesca Annese, Giulia Fontò, Alessandra Stasi, Francesco Pesce, Silvio Tafuri, Vincenzo Di Leo, Loreto Gesualdo

**Author notes:** **Corresponding author:** Fabio Sallustio, PhD; Department of Precision and Regenerative Medicine and Ionian Area (DIMEPRE-J), University of Bari, Piazza G. Cesare,11 70124 Bari, Italy.

## Abstract

Immunoglobulin A nephropathy (IgAN) is the most frequent primary glomerulonephritis characterized by the presence of IgA immune complexes in the glomeruli. Recently, a multihit model has been proposed to describe the pathogenesis of IgAN, but it is believed that further predisposing factors are present, including immunological, genetic and environmental factors. Newly, the role of IL-6 in pathogenesis is becoming increasingly important but reason why levels of IL-6 are elevated in IgAN patients is not well understood.

One attainable hypothesis on high levels of IL-6 in IgAN comes out from our recent whole genome DNA methylation screening in IgAN patients, that identified, among others, a hypermethylated region comprising Vault RNA 2-1 (VTRNA2-1), a non-coding RNA.

Here we confirm that VTRNA2-1 is low expressed in IgAN subjects compared to HS and we found that also in transplanted IgAN patients (TP-IgAN), compared to non-IgAN transplanted patients (TP), the VTRNA2-1 transcript was expressed at level very low. We found that in IgAN patients with downregulated VTRNA2-1, PKR is overactivated, coherently with the role of the VTRNA2-1 that binds to PKR and inhibits its phosphorylation. The loss of the VTRNA2-1 natural restrain caused the activation of CREB by PKR, a classical cAMP-inducible CRE-binding factor interacting with a region of the IL-6 promoter and leading to IL-6 production, both in IgAN and in TP-IgAN patients. PKR is normally activated by bacterial and viral RNA and we found that both the RNA poly(I:C), the and the COVID-19 RNA-vaccine stimulation significantly increase the IL-6 levels in PBMCs from HS but had an opposite effect in those from IgAN patients.

In conclusion, the discovery of the upregulated VTRNA2-1/PKR/CREB/IL-6 pathway in IgAN patients may provide novel approach to treat the disease and may be useful for development of precision nephrology and personalized therapy, possibly by checking the VTRNA2-1 methylation level in IgAN patients.

## INTRODUCTION

Immunoglobulin A nephropathy (IgAN), also known as Berger’s disease, is the most frequent primary glomerulonephritis characterized by the presence of IgA immune complexes in the glomeruli (1). It generally appears in the second and third decade of life, has a higher incidence in males and it is more common in whites than in blacks with a higher prevalence in Asians than Caucasians(1).

Recently, a multihit model has been proposed to describe the pathogenesis of IgAN. In this model, the first hit is given by the hypersecretion of deglycosylated IgA (Gd-IgA1)(2). These Gd-IgA1 are recognized and attacked by autoantibodies (second hit) and this process leads to the formation of circulating immune complexes (third hit), some of which are deposited at the mesangial level in the glomeruli(2). Kidney damage resulting from the deposition of the cells is characterized by local inflammation, complement activation, cell proliferation, and finally fibrosis(3).

Moreover, in humans, the gut-associated lymphoid tissue (GALT) is the primary source of IgA, therefore the pathogenesis of IgAN is related to gut homeostasis. Indeed, the intestinal–renal axis is important in Berger’s glomerulonephritis, where several factors (e.g. genetics(4,5), infections(6,7), and food antigens (8) may play a role in the disease complex pathogenesis and provide novel therapeutic targets to modify disease evolution.

These models explain the pathogenesis of IgAN caused by the production of aberrant IgA, but it is believed that further predisposing factors are present, including immunological, genetic, environmental, or nutritional factors that can influence the pathogenesis and that could be useful for development of precision nephrology and personalized therapy.

We have recently showed that IgAN patients have a higher frequency of intestinal-activated B cells than healthy subjects (HS). IgAN patients showed greater BAFF cytokine blood levels, which were linked to higher levels of five microbiota metabolites, and high APRIL cytokine serum levels. In comparison to HS, IgAN individuals had a larger number of circulating gut-homing (CCR9 β7 integrin) regulatory B cells, memory B cells, and IgA memory B cells(9).

Moreover, several studies have shown the involvement of the IL-6 pathway in IgAN. Interleukin-6 is essential for glomerular IgA deposition and the development of renal pathology in Cd37-deficient mice (10). In addition, the proliferation-inducing ligand (APRIL) and IL-6 are involved in the overproduction of aberrantly glycosylated IgA. In mice, the APRIL silencing blocked the overproduction of Gd-IgA1 induced by IL-6 and, conversely, neutralizing IL-6, the production of Gd-IgA1 was reduced(11). Interestingly, the mycotoxins deoxynivalenol (DON) prolonged exposure cause the expansion of IgA secreting B cells by activating macrophages and T cells. In mice this stimuation results in the early stages of human IgA nephropathy(12,13) and the overproduction of inflammatory interleukins such as IL-6(14), which in turn induces upregulation of IgA.

However, to date, the biological mechanisms leading to the elevate IL6 levels in IgAN patients are not clear. In a recent study, a whole-genome screening was performed for DNA methylation in CD4^+^ T cells from IgAN patients, identifying three regions with altered methylation capable of influencing the gene expression of genes involved in cell response and proliferation T CD4^+^(15). In particular, a hypermethylated region was identified comprising Vault RNA 2-1 (VTRNA2-1), a non-coding RNA also known as the precursor of miR-886 (Pre-mi-RNA).

Here we studied the VTRNA2-1/PKR/CREB/IL-6 pathway showing that it is upregulated in IgAN patients and that it is responsible for the elevated IL-6 levels characterizing the disease, thus providing for the first time an explanation on the abnormal levels of this cytokine. We found that this pathway is epigenetically controlled by VTRNA2-1 and modulated by bacterial and viral infections. It can explain both the high levels of IL-6 and the correlation to mucosal infections.

Finally, we showed that the drug imoxin, targeting this pathway, can reduce IL-6 secretion by PBMCs of IgAN patients providing a new possible approach to treat the disease.

## RESULTS

### VTRNA2-1 is downregulated in IgAN patients

Recently, we showed that the non-coding RNA VTRNA2-1 was epigenetically downregulated in IgAN patients(15). Since we hypothesize that it may be implicated in the pathway for the secretion of IL-6, we further confirm the involvement of VTRNA2-1 in IgAN studying its expression in PBMCs isolated from IgAN patients and healthy subjects.

VTRNA2-1 was strongly downregulated in IgAN patients compared to healthy subjects showing a decrease of 30 fold (p <0.05, Figure 1A). Moreover, recent clinical observations have shown that IL-6 production is implicated in renal allograft rejection(16,17) and that cumulative risk of IgA nephropathy recurrence increased after transplant and was associated with a 3.7-fold greater risk of graft loss (17).

**Figure 1.**
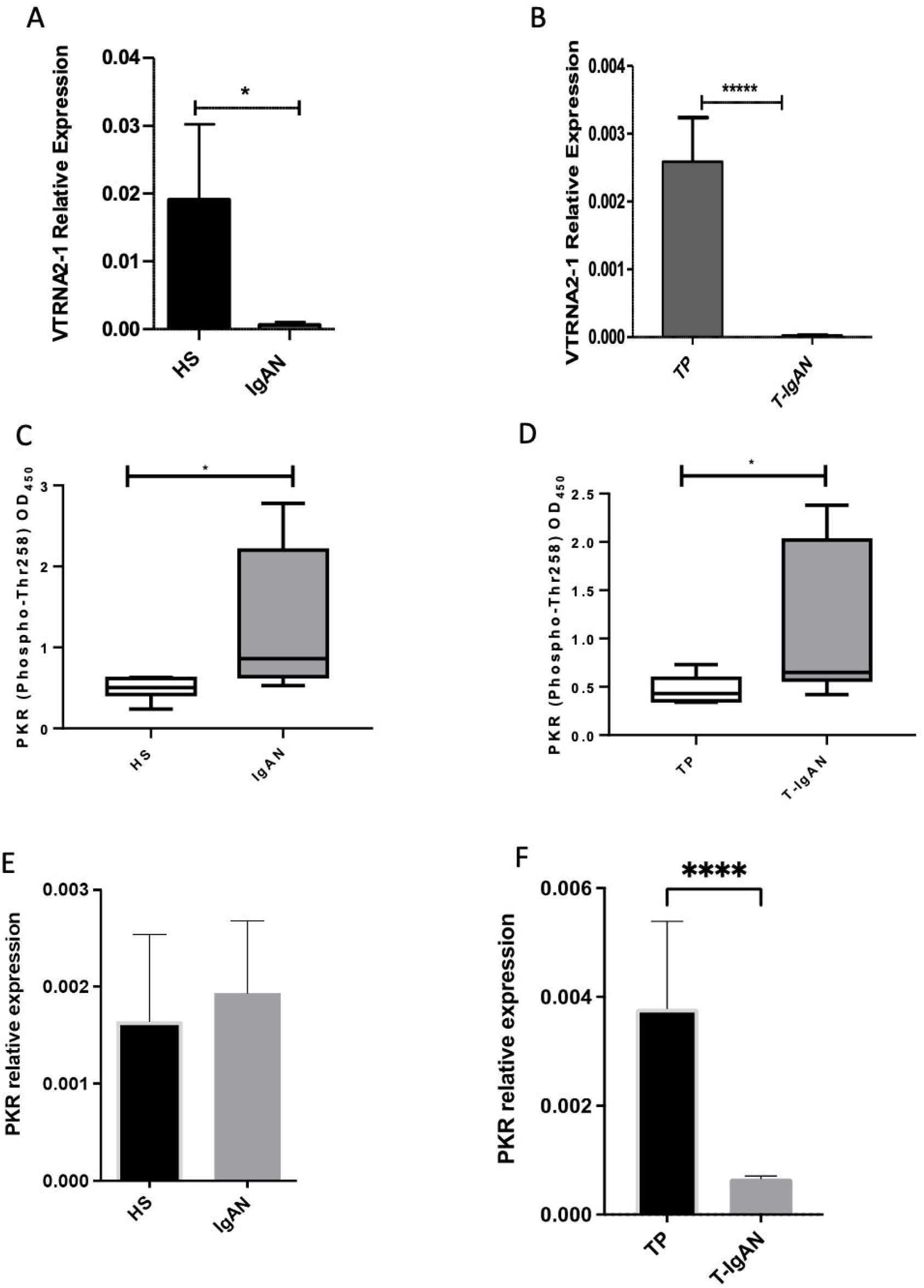
IgAN patients showed extremely low VTRNA2-1 levels and high levels of phosphorylated PKR. (A)Real-time PCR of VTRNA2-1 transcript differentially expressed in PBMCs of IgAN and HS. (B) Real-time PCR of VTRNA2-1 transcript differentially expressed in PBMCs of T-IgAN and TP. (C) ELISA assay showed significantly increased level of phosphorylated PKR in IgAN patients respect to healthy subjects (HS). (D) A strong increase in phospho-PKR levels was found in T-IgAN patients compared to TP group. (E) Gene expression of PKR in IgAN patients and HS. (F) Gene expression of PKR in TP and T-IgAN patients. Data are representative of 8 independent experiments and are expressed as mean ± SEM, (* p-value < 0.05, ****p-value <0.001).

We therefore analysed the expression of VTRNA2-1 in PBMCs isolated from IgAN patients with kidney transplant (T-IgAN) and in patients non-IgAN with kidney transplant (TP). Also in this case, we found that the VTRNA2-1 was extremely downregulated in transplanted IgAN patients compared to their controls (p<0.0001, fold change=100; Figure 1B), confirming that this modulation is a characteristic of IgAN disease.

### Downregulation of VTRNA2-1 In IgAN patients lead to the increase in phosphorilated PKR

VTRNA2-1 can inhibit PKR, an interferon-inducible and double-stranded RNA (dsRNA) dependent kinase, by binding to this protein and preventing its auto-phosphorylation(18). Thus, we studied whether in the IgAN patients with low levels of VTRNA2-1 the correspective upregulation of the PKR signalling was present. We measured the levels of the phosphorilated (pPKR) and of the total PKR and found that in IgAN patients the level of pPKR protein normalized on the total PKR was significantly increased compared to HS (FC 2.6; p<0.05; Fig. 1C). At the same manner, also in T-IgAN we found the doubling of pPKR levels compared to TP (FC 2, p<0.05; Fig. 1D).

To confirm that the incerase in pPKR levels was due to the auto-phosphorylation induced by the lack of VTRNA2-1 and not by a different mechanism regulating the gene expression, we checked the PKR mRNA levels in IgAN patients and in HS. We found no significant differences between the two group (Figure 1E). However, in T-IgAN compared to TP we found rather a decrease of the PKR transcripts (Figure 1F, P<0.0001), making data of the pPKR increase even more meaningful.

### Phosphorylation of PKR corresponds to CREB activation in IgAN patients

In mice pPKR induced the Cyclic adenosine monophosphate (AMP) response element-binding protein (CREB) activation by increasing its phosphorilation (12). We studied whether in IgAN patients CREB phosphorilation levels were higher compared to HS. A significant 3-fold increase of pCREB levels was found in IgAN patients compared to HS (p<0.01; Fig. 2A). Also in T-IgAN pCREB levels were significantly increase compared to TP, even if to less extent (1.67 fold, p<0.01; Fig. 2B). However, no significant difference in CREB gene expression was present among IgAN patients and HS (Figure 2C), nor in TX compared to T-IgAN (Fig. 2D), corroborating the data of CREB phosphorilation in IgAN patients.

**Figure 2.**
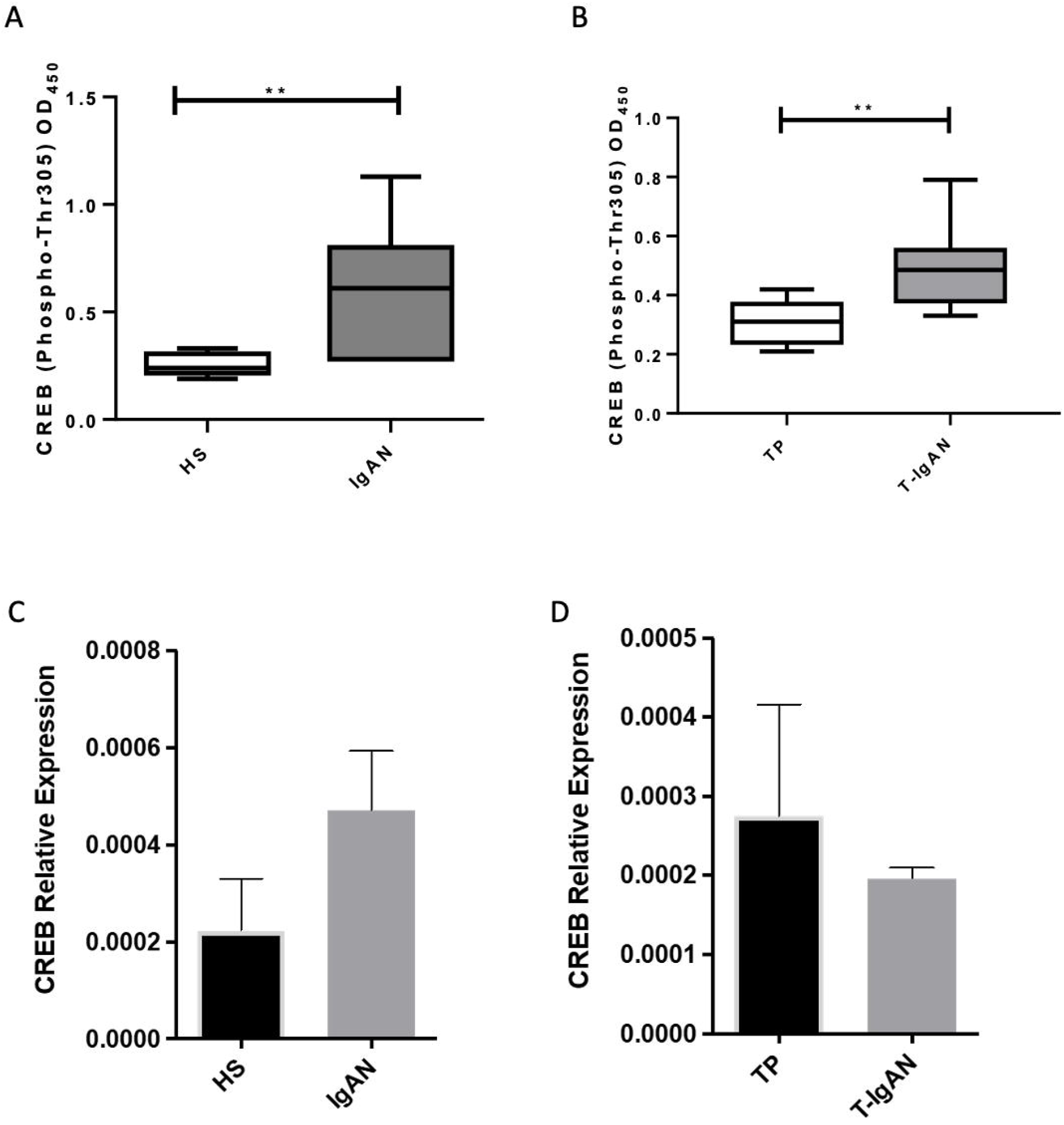
IgAN patients with low VTRNA2-1 showed elevated levels of phosphorylated CREB. (A) The assay revealed systemic activation of CREB (phospho-Thr305-CREB) in IgAN group compared to healthy subjects (HS). (B) CREB phosphorylation was significantly increased in T-IgAN patients compared to transplanted patients without IgAN disease. (C) Gene expression of CREB in IgAN patients and HS. (D) Gene expression of CREB in TP and T-IgAN patients. Data are representative of 8 independent experiments (means ±SEM; **p-value <0.01).

### CREB activation by phosphorylation leads to IL-6 increase in IgAN patients

Since several studies showed that CREB activation may lead to IL-6 expression(19–23), we analyzed the IL-6 levels secreted by PBMCs in our IgAN patient groups. IL-6 levels showed a statistically significant increase in IgAN patients compared to HS (p<0.05; Fig. 3A). In T-IgAN compared to TP a tendence in increased levels of IL-6 was found, even if it is not significant (Fig. 3B). Both the levels of pCREB and pPKR significantly correlated with IL-6 levels in IgAN patients (r=0.97, p= 0.0006 and r=0.89, p=0.0064, respectively), indicating that the high levels of this cytokine are due to the VTRNA2-1/pPKR/pCREB signalling epigenetically regulated (Figure 3C-D). Both pCREB and IL-6 levels significanlty increased in IgAN patients and not in minimal change diseases patients (MCD) compared to healthy subjects, confirming that the PKR/CREB/IL-6 pathway is hyperactivated specifically in IgAN (Figure 3E-F).

**Figure 3.**
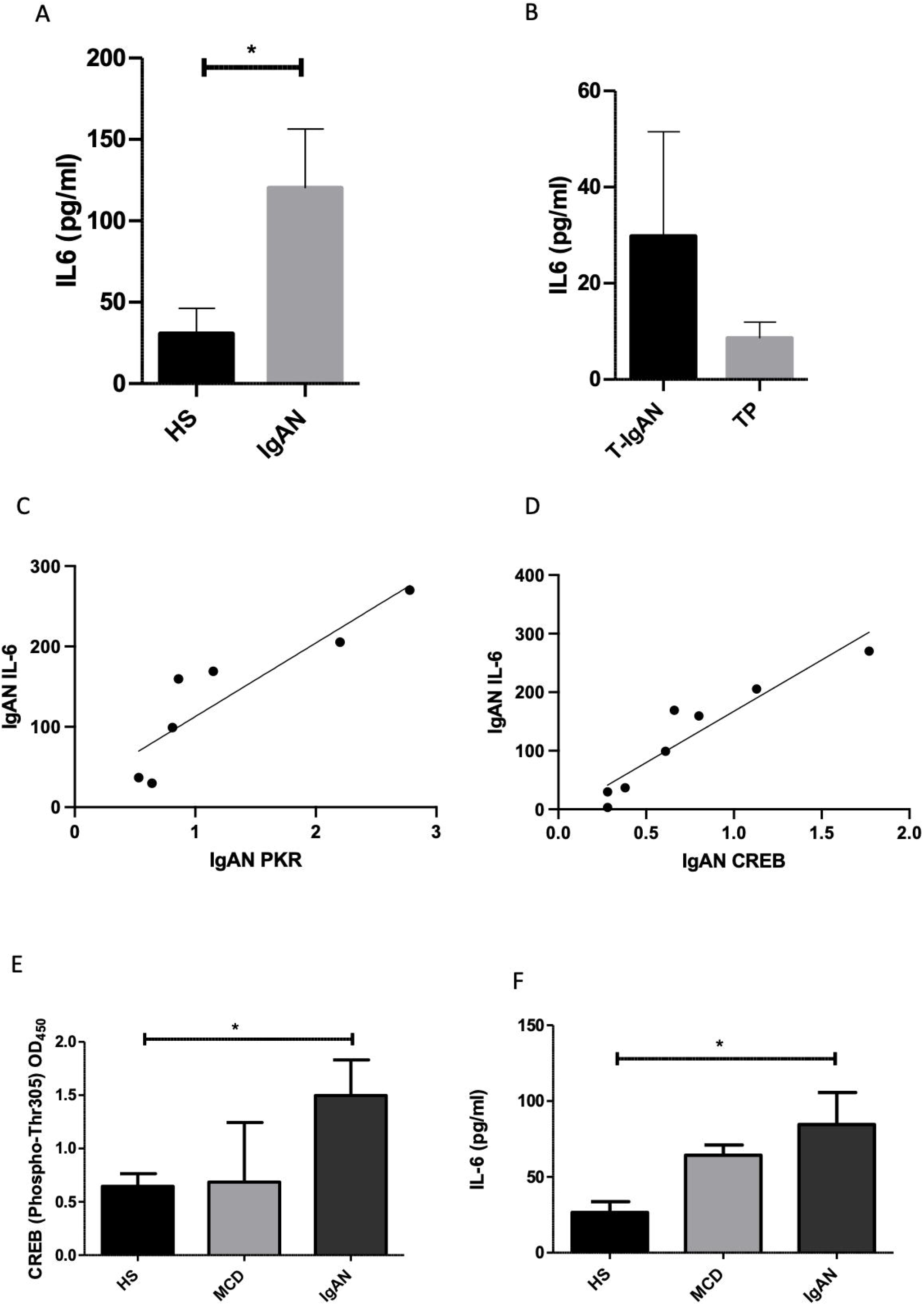
VTRNA2-1/PKR/CREB activated pathway leads to elevated IL-6 secretion in IgAN patients. (A) ELISA assay showed significant increased levels of IL-6 in IgAN patients with VTRNA2-1/PKR/CREB/IL-6 activated pathway respect to healthy group. (B) An IL-6 increase was found in T-IgAN patients with VTRNA2-1/PKR/CREB activated pathway compared to TP group, even if not statistically significant. Both the levels of pPKR (C) and pCREB (D) significantly correlated with IL-6 levels in IgAN patients (r=0.97, p= 0.0006 and r=0.89, p=0.0064, respectively). Data are representative of 8 independent experiments. (E) CREB phosphorylation was significantly increased in IgAN patients but not in MCD patients compared to HS. (F) IL-6 levels were significantly increased in IgAN patients but not in MCD patients compared to HS. Data are expressed as means ±SEM; * p-value < 0.05.

### PKR inhibitor Imoxin decrease IL-6 secretion by PBMCs from IgAN patients

To further validate the dependence of IL-6 secretion from PKR activation we investigated the effect of the PKR inhibitor imoxin, named also imidazol or C16, as a blocking agent for the pCREB activation and IL-6 secretion. Results showed that both CREB phosphorilation and IL6 secretion in IgAN patient PBMCs were significantly reduced by 1 μM imoxin stimulation for 48 hours (Figure 4 A-B, p<0.05 and p<0.01, respectively). However, imoxin stimulation of PBMC from MCD patients did not decrease nor CREB activation nor IL-6 levels (Figure 4C-D). Since PKR is a selective inhibitor of PKR, these data confirmed that the PKR/CREB/IL-6 pathway is specifically activated in IgAN patients.

**Figure 4.**
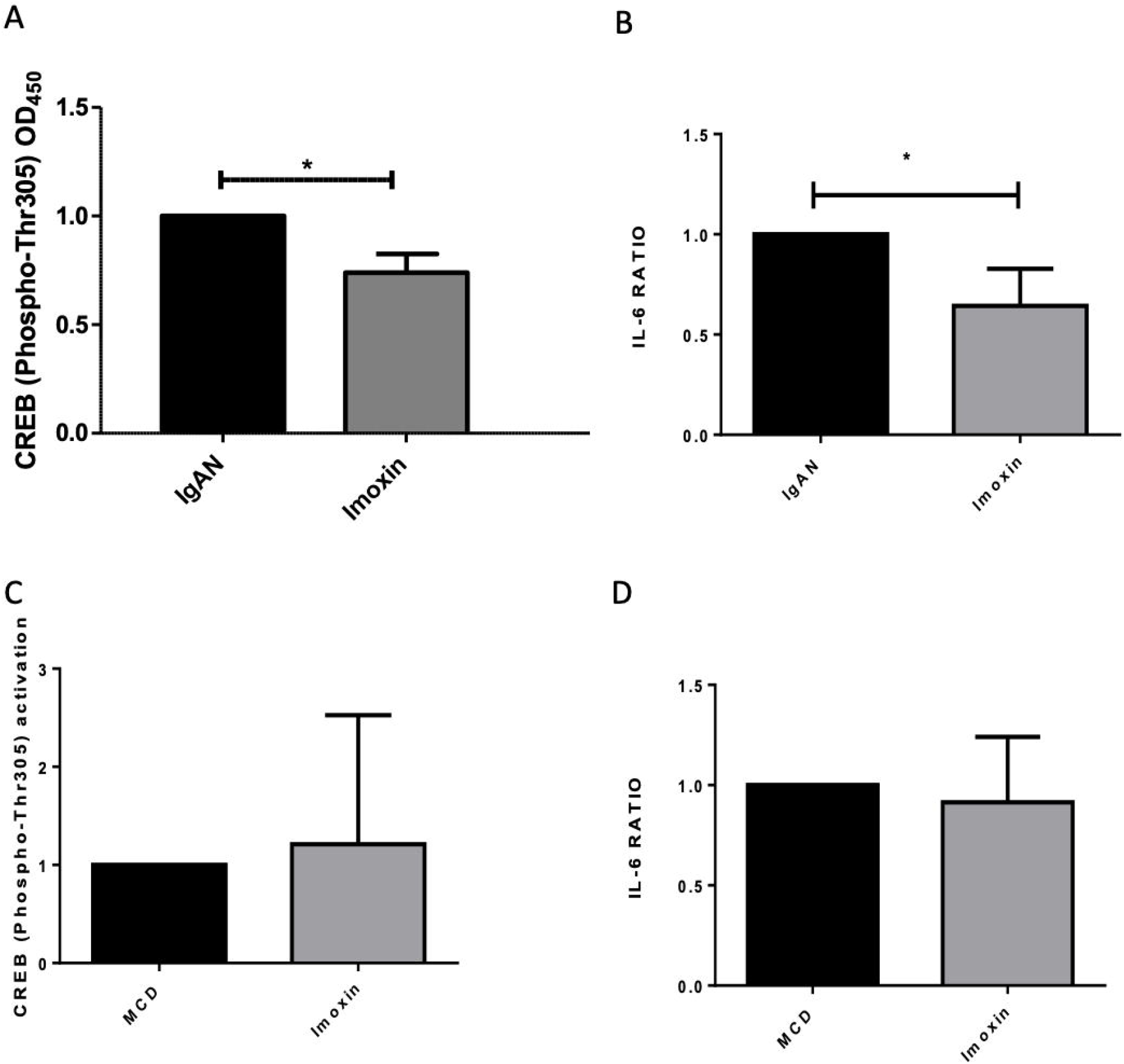
PKR Inhibitor Imoxin decrease CREB activation and IL-6 secretion in PBMCs from IgAN patients. (A) ELISA assay showing that imoxin significantly decrease the CREB activation in PBMCS from IgAN patients. (B) ELISA assay showing that imoxin significantly decrease the IL-6 secretion in PBMCS from IgAN patients. (C) ELISA assay showing that imoxin did not decrease CREB activation in PBMCS from MCD patients. (D) ELISA assay showing that imoxin did not decrease IL-6 secretion in PBMCS from MCD patients. Data are expressed as means ±SEM; * p-value < 0.05.

### Double and single strand RNA or COVID vaccine stimulation increase IL-6 secretion in PBMCs from HS and has an opposite effect in PBMCs from IgAN patients

The biological effect of triggering the PKR/CREB/IL-6 pathway on PBMCs from IgAN patients and HS was studied. Since PKR can be activated by viral or bacterial RNA (24,25) (26,27), we transfected lymphocytes with synthetic polyinosinic:polycytidylic acid (poly(I:C)), which mimics dsRNA or with the RNA vaccine against SARS-COV2, the renowned virus causing COVID-19. After 48h of treatment, PBMCs from HS treated with Poly(I:C) showed a 3-fold increase in the IL-6 ratio as compared to the untreated condition, while a more modest increase in IL-6 secretion was detected after vaccine treatment. Nonetheless, both dsRNA and ssRNA trigger IL-6 production in healthy subjects (Figure 5A-B, p<0.05). Unexpectedly, when we extended our investigation to IgAN patients, we found that both the Poly(I:C) and the vaccine stimulation had an opposite effect compared to the HS stimulation leading to a decreased IL-6 secretion (Figure 5C-D, p<0.05). The RNA inhibitor effects in IgAN patients was confirmed also transfecting the PBMC directly with the SARS-CoV-2 RNA (Figure 5E, p<0.001). The restraint of the CREB phosphorilation following RNA stimulation in PBMC from IgAN patients confirmed the different PKR/CREB/IL-6 pathway control in IgAN patients compared to HS (Figure 5F-H). Both the RNA Covid and the vaccine stimulation induced a significant decrease od CREB phosphorilation (Figure 5F-E, p<0.05 and p<0.01, respectively); however the Poly(I:C) did not change the CREB activation (Figure 5H).

**Figure 5.**
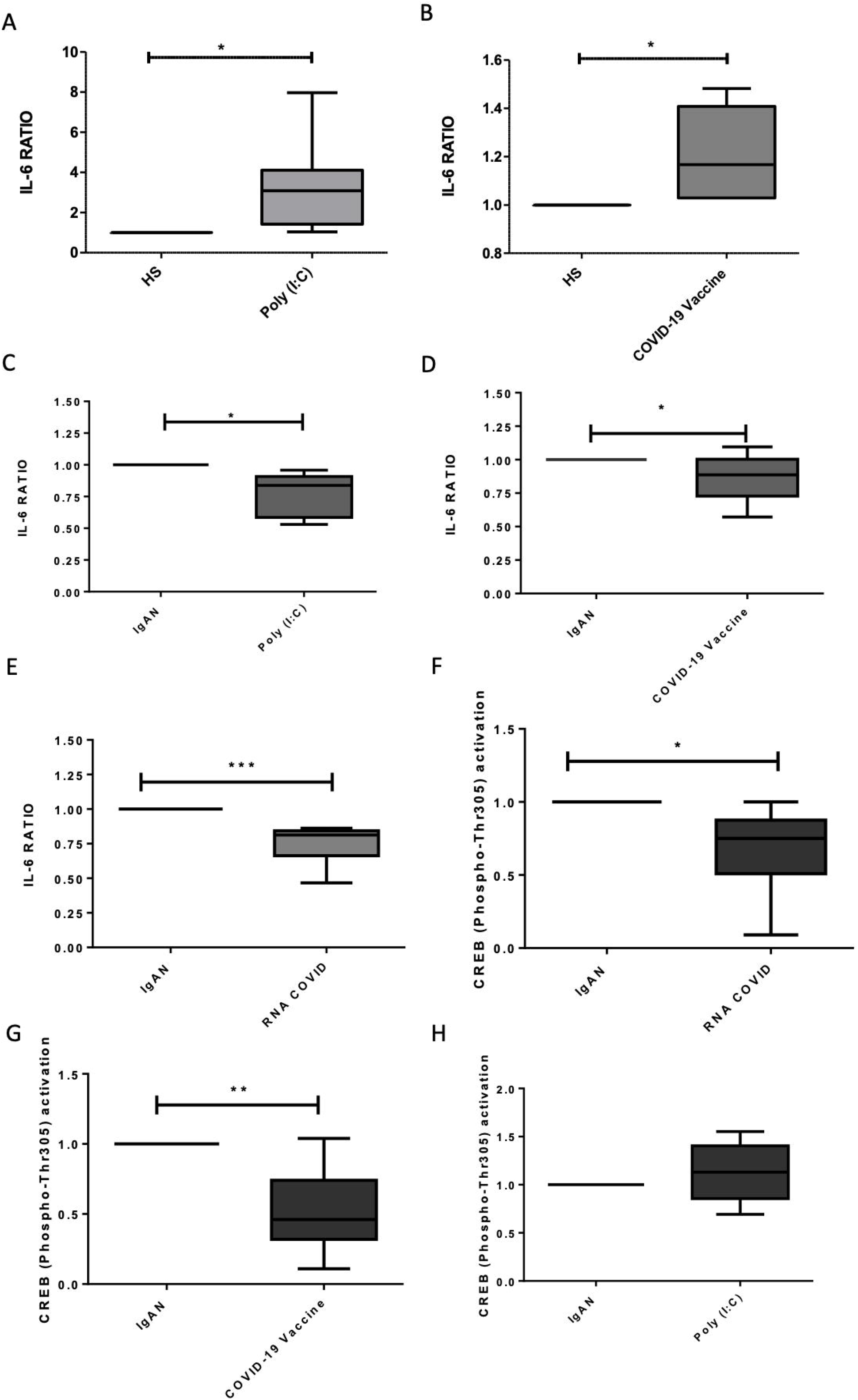
Effect of poly(I:C), COVID vaccine, and SARS-CoV-2 RNA stimulation in IL-6 secretion in PBMCs from HS and IgAN patients. (A) ELISA assay showing that the poly(I:C) stimulation increased IL-6 secretion in PBMCs from HS. (B) ELISA assay showing that Covid 19 vaccine stimulation increased IL-6 secretion in PBMCs from HS. (C) ELISA assay showing that the poly(I:C) stimulation decreased IL-6 secretion in PBMCs from IgAN patients. (D) ELISA assay showing that Covid 19 vaccine stimulation decreased IL-6 secretion in PBMCs from IgAN patients. (E) ELISA assay showing that Covid RNA stimulation decreased IL-6 secretion in PBMCs from IgAN patients. (F) ELISA assay showing that Covid RNA stimulation decreased CREB activation in PBMCs from IgAN patients. (G) ELISA assay showing that Covid 19 vaccine stimulation decreased CREB activation in PBMCs from IgAN patients. (H) ELISA assay showing that the poly(I:C) stimulation did not change CREB activation in PBMCs from IgAN patients. Data are expressed as means ±SEM; * p-value < 0.05; **p-value <0.01.

**Figure 6.**
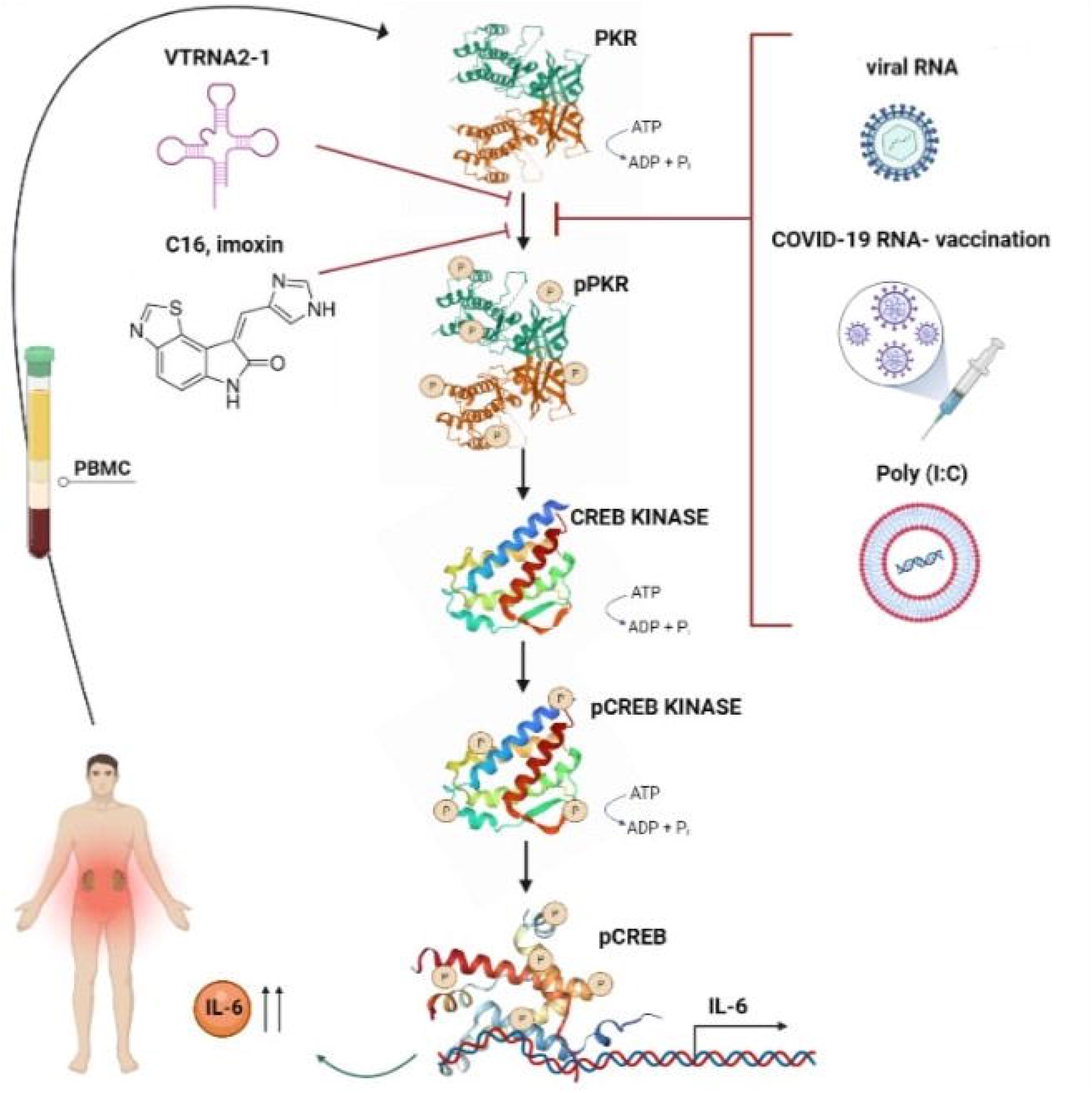
The involvment of PKR/CREB/IL-6 pathway in IgAn patients and its modulation. In IgAN patients the PKR/CREB/IL-6 pathway is overexpressed and lead to the IL-6 secretion by patient PBMCs. The pathway is modulated by the non-coding RNA VTRNA2-1 that is downregulated (because it is hypermetilated) and therefore can not inhibit PKR phosphorylation. As a consequence, PKR is hyperphosphorylated and, in turn, actives phosphorylation of CREB kinase. Posho CREB (pCREB) interacts with a region of the IL-6 promoter and leads to high levels of IL-6 production. PKR is also induced by bacterial and viral RNA, thus PBMC transfection with synthetic polyinosinic:polycytidylic acid (poly(I:C)) and COVID-19 vaccine normally increase IL-6 levels in HS. Instead, in IgAN patients this stimulation had an opposite effect compared to the HS stimulation leading to a decreased IL-6 secretion. Moreocer, the drug Imoxin is a PKR inhibitor, that prevents PKR phosphorylation and as a consequence inhibits the PKR/CREB/IL-6 pathway, with a significantly decrease of IL-6 secretion.

## DISCUSSION

The exact immunopathogenic mechanisms underlying IgAN are poorly understood. Very recently the role of Il-6 in IgAN pathogenesis is becoming increasingly important(28–32), even if the reason why levels of IL-6 are elevated in IgAN patients is not well understood.

It selectively increases production of galactose-deficient IgA1 in IgA1-secreting cells from patients with IgA nephropathy(28,29) and IL6-/-mice displayed no glomerular IgA deposition and were protected from exacerbated renal failure following lipopolysaccharide treatment(31). Interestingly, IL-6 signalling seems to be involved in this complex intestinal immune network particularly in mediating the production of Gd-IgA(10,33).

One attainable hypothesis on high levels of IL-6 in IgAN comes out from our recent whole genome DNA methylation screening in IgAN patients, that identified, among others, three regions with altered DNA methylation capable of influencing the expression of genes involved in the response and proliferation of T and B cells (15). We identified a hypermethylated region encompassing Vault RNA 2-1 (VTRNA2-1), a non-coding RNA also known as precursor of miR-886 (pre-mi-RNA). Consistently, the VTRNA2-1 expression was found down-regulated in IgAN patients (15).

Here we confirm that VTRNA2-1 is lower expressed in IgAN subjects compared to HS and we found that also in transplanted IgAN patients compared to non IgAN transplanted patients the VTRNA2-1 transcript was expressed at very low levels. However, the RNA product does not function as a vault or microRNA; rather, it acts as a direct inhibitor of protein kinase R (also known as eukaryotic translation initiation factor 2-alpha kinase 2, EIF2AK2), and thereby plays an important role in the regulation of cell growth. This gene is nearby a differentially methylated region (DMR), it is imprinted and may show allele-specific expression. VTRNA2-1 is a metastable epiallele with accumulating evidence that methylation at this region is heritable, modifiable, and associated with disease including risk and progression of cancer(34).

We found that, in IgAN patients with downregulated VTRNA2-1, PKR is overactivated, coherently with the role of VTRNA2-1 that binds to PKR and inhibits its phosphorylation (35). The loss of this natural restrain causes the activation of CREB, a classical cAMP-inducible CRE-binding factor interacting with a region of the IL-6 promoter (that is known as CRE-like sequence), leading to IL-6 production (19–23).

The discovery of the upregulation of the PKR/CREB/IL-6 in IgAN patients is very suggestive. PKR is normally activated by double-strand bacterial(24,25) and single-strand viral (26,27) RNA and has long been recognized as a key mediator of the innate immunity response to viral infection. Expression of latent PKR is induced by interferon and it is activated upon binding to viral RNA containing duplex regions to undergo autophosphorylation.

We expected to find a further activation of PKR and of the downstream pathway following the PBMC stimulation by RNA, simulating a bacterial or viral infection, or also the COVID vaccination. Instead, this mechanism works in HS but not in IgAN patients. Our data cannot explain the reasons, but we can hypothesize that, in subjects with this pathway already hyperactivated, the further triggering led to an inhibition rather than an extra activation. This hypothesis is supported by studies showing that increasing nucleic acid concentration leads to PKR inhibition, likely because PKR monomers are diluted out on separate dsRNA molecules and have therefore decreased ability to dimerize (36–38) However, the epigenetic hyperactivation of the PKR/CREB/IL-6 pathway may contribute to the IgAN onset together to the other determinants. The data showing that in IgAN patients the PKR signalling is already activated due to their epigenetic background could imply a sort of balance that may be perturbed by mucosal infections leading to clinical manifestation of hematuria following upper respiratory tract microbes challenge(39,40). Interestingly, very recently it has been shown that PKR is regulated by adenovirus-associated noncoding RNA that functions by binding PKR but not inducing activation, thereby inhibiting the antiviral response (41). This kind of mechanism, driven by the epigenetic silencing of VTRNA2-1, may therefore explain both the high levels of IL-6, and recent data showing microbiota involvment in IgAN (9) (42) (43).

The PKR/CREB/IL-6 pathway may be very important also in the setting of renal transplantation. We found that this pathway is upregulated also in IgAN transplanted patients. Recent studies showed that the cumulative risk of IgA nephropathy recurrence increases after transplant and is associated with a 3.7-fold greater risk of graft loss(44). Besides, evidence suggests that IL-6 may play an important role in donor-specific antibodies generation and chronic active antibody-mediated rejection and that the treatment with an anti-IL-6 receptor monoclonal antibody may represent a novel approach for chronic antibody-mediated rejection and transplant glomerulopathy, stabilizing allograft function and extend patient lives (17).

Finally, we showed that the IL-6 secretion can be reduced by the PKR inhibitor imoxin (Figure 7). This drug also known as C16 or imidazole, has shown beneficial effects on cell cultures and in vivo in animal models for numerous conditions, such as improvement in inflammation, oxidative stress, diabetes, suppression of tumor proliferation and in the control of hypertension (45). However, in light of our results, it may be considered as a possible therapeutic drug also in IgAN. Interestingly, in mice dietary exposure to the common foodborne mycotoxin deoxynivalenol (DON) upregulates serum immunoglobulin A (IgA) and IL-6 miming IgAN and activating PKR/CREB/IL-6 pathway. The dietary omega-3 fatty acids can invert these processes and ameliorate DON-induced IgA nephropathy (12,13).

Further studies will be needed to address this point and the weight of the involvement of the VTRNA2-1/PKR/CREB/IL-6 pathway in IgAN, also increasing the number patients to study. Moreover, we analysed only Caucasian patients, but the results should be confirmed in other ethnicities.

In conclusion, the discovery of the upregulated VTRNA2-1/PKR/CREB/IL-6 pathway in IgAN patients open new perspectives in the study of the disease onset and development and may provide novel approach to treat the disease, including the use of imoxin as a drug in IgAN patients showing the PKR, or the entire pathway, hyperactivated. Our results suggest that, in the future, a screening for VTRNA2-1 methylation/expression may be useful for development of precision nephrology and personalized therapy for the IgAN disease.

## MATERIALS AND METHODS

### Study design and patients

The study was carried out in accordance with the Helsinki Declaration and the European Guidelines for Good Clinical Practice and approved by our institutional ethics review. Five groups of Caucasian volunteers were included in the study after providing their written informed consent: 34 primary biopsy-proven IgAN patients, 22 IgAN subjects with kidney transplant (T-IgAN), 14 patients transplanted for causes other than IgAN (TP), 11 HS without known diseases and 5 controls with non-IgA glomerulonephritis (minimal change disease), matched to cases by age and gender. The main clinical features of enrolled patients and HSs included in the study are summarized in **IgAN** All patients were enrolled before receiving drug administrations. No significant difference in age and sex distribution was observed among the groups.

PBMCs were isolated by gradient centrifugation with Ficoll-Hypaque (Euroclone, Italy) from heparinized venous blood from patients and HS. PBMCs were cultured in RPMI 1640 (Euroclone) supplemented with 10% Fetal Bovine Serum (Euroclone), 100U/mL Penicillin/Streptomycin (Euroclone), 4mMGlutamine (Euroclone), 10 mM Hepes (Sigma), 0.1 mM non-essential amino acids (Euroclone), 1mM sodium pyruvate and 50 U/mL, rhIL-2.

### RNA extraction and real time-PCR expression of VTRNA2-1, CREB and PKR

Total RNA were isolated from PBMCs of IgAN, T-IgAN, TP and HS using RNeasy Mini Kit (Qiagen, Hilden, Germany) according to the manufacturer’s instruction and quantified by NanoDrop One Spectrophotometer (thermoFisher Scientific). Total RNA was retro-transcribed using the miScript II RT Kit with iScript cDNA Synthesis Kit (Bio-Rad, Hercules, CA, USA)following the manufacturer’s instructions.

Real Time-PCR was performed on a StepOne Plus instrument (Applied Biosystems) by using VTRNA2-1, PKR and CREB primers (Integrated DNA Technologies, Coralville, IA,USA) GAPDH gene amplification was used as a reference standard to normalize the target signal. Real Time-PCR was performed in triplicate, and relative expression was calculated using the 2−ΔC_t_ method.

### pPKR/pCREB/IL-6 pathway ELISA quantification

Colorimetric Cell-Based ELISA assays were used to detect the levels of CREB/pCREB (CREB (Phospho-Ser142) Aviva Systems Biology, USA), and PKR/pPKR (PKR (Phospho-Thr258) Aviva Systems Biology, USA).

Briefly, PBMCs of HS, IgAN, T-IgAN and TP subjects were freshly isolated and seeded overnight with RPMI medium at 37°C, 5% CO_2_, in a 96-Well Cell Culture Clear-Bottom Microplate. The ELISA assay protocols were executed following manufacturer instructions.

To determine IL-6 levels, PBMCs of HS, IgAN, T-IgAN and TP subjects, were freshly isolated and seeded in a 96-Well Cell Culture Microplate supplemented with RPMI medium at 37°C, 5% CO_2_. After 48 h of culture, cells were collected and, centrifuged at 230 rpm for 15 minutes. Supernatant were recovered and analysed using Human IL-6 Quantikine ELISA Kit (R&D Systems, USA), following manufacturer’s protocol.

Optical densities were determined using a microplate reader (programmable MPT reader model DV 990BV6; GDV, Italy) set to 450 nm.

### Transfection experiments

PBMCs of HS and IgAN patients were seeded in 6-well plates at a density of 2 × 10^6^ cells/cm^2^ and were incubated with specific medium. Xfect Transfection Reagent (Clontech,Takara bio, France) was used to transfect 20 ug of Poly (I:C) (InvivoGen, Europe), a synthetic analogue of dsRNA, in PBMC cells, according to manufacturer’s instructions. Imoxin (Chem Cruz) was added to PBMCs at a concentration of 1 μM. After two days of incubation with Poly (I:C) and/or Imoxin the secretion of IL-6 levels by PBMCs were measured in the cell culture supernatants.

### Statistical Analysis

Statistical analyses were performed using the Student’s t-test, as appropriate. A p-value < 0.05 was considered significant. Data are expressed as means ± SEM.

## Data Availability

All data produced in the present work are contained in the manuscript

## Notes

### Competing Interest Statement

The authors have declared no competing interest.

### Funding Statement

This study was funded by University of Bari

### Author Declarations

Ethics committee of University Polyclinic Hospital of Bari gave ethical approval for this work (Prot. N. 606)

### Summary of Updates

New experiments added

